# Association between myopia and peripapillary hyperreflective ovoid mass-like structures in children

**DOI:** 10.1101/19001180

**Authors:** In Jeong Lyu, Kyung-Ah Park, Sei Yeul Oh

## Abstract

**Purpose:** To investigate the characteristics of children with peripapillary hyperreflective ovoid mass-like structures (PHOMS) and evaluate the risk factors associated with PHOMS.

**Methods:** This study included 132 eyes of 66 children with PHOMS and 92 eyes of 46 children without PHOMS (controls) who were assessed by disc enhanced depth image spectral-domain optical coherence tomography (OCT). Univariable and multivariable logistic analyses were performed to evaluate risk factors associated with the presence of PHOMS.

**Results:** Among the 66 children with PHOMS, 53 patients (80.3%) had bilateral and 13 patients (19.7%) had unilateral PHOMS. The mean age of the PHOMS group was 11.7 ± 2.6 years and 11.4 ± 3.1 years in the control group. Mean spherical equivalent (SE) by cycloplegic refraction was −3.13 ± 1.87 diopters (D) in the PHOMS group and −0.95 ± 2.65 D in the control group. Mean astigmatism was 0.67 ± 0.89 D and 0.88 ± 1.02 D in the PHOMS group and the control group, respectively. Mean disc size was 1735 ± 153 μm in the PHOMS group and 1741 ± 190 μm in the control group. All eyes in PHOMS group had myopia of −0.50 D or less, except for an eye with +1.00 D. According to the univariable (odds ratio [OR] 1.59, *P* < 0.001) and multivariable (OR 2.00, *P* < 0.001) logistic regression analyses, SE decreased by 1 D was significantly associated with PHOMS.

**Conclusions:** PHOMS is associated with myopic shift in children. Optic disc tilt may be a mediator between myopia and PHOMS.

## Introduction

Optic disc drusen (ODD) are diseases of optic nerve head with acellular hyaline depositions (Pollack & Becker 1962; Friedman et al. 1977; Tso 1981). It has been suggested that ODD be categorized as buried or superficial according to the morphologic findings on funduscopic examination and optical coherence tomography (OCT) images (Auw-Haedrich et al. 2002; Lee et al. 2013). Buried drusen are reported as the main feature of ODD in younger patients and more difficult to differentiate from papilledema than superficial drusen (Auw-Haedrich et al. 2002; Lee et al. 2013; Casado et al. 2014). However, OCT findings of buried ODD have been the subject of recent debate (Lee et al. 2018b; Malmqvist et al. 2018a, b). Optic Disc Drusen Studies Consortium suggested that hyperreflective mass-like structures on OCT, which have been described as configurations of buried drusen (Lee et al. 2011; Gili et al. 2013; Lee et al. 2013; Bassi & Mohana 2014; Lee et al. 2018a), are not true ODD (Malmqvist et al. 2018b). The authors labeled this finding as peripapillary hyperreflective ovoid mass-like structures (PHOMS) and suspected that these structures result from the herniation of distended axons into the peripapillary retina. The ODDS has set definition of ODD on OCT as hyporeflective structures with full or partial hyper-reflective margin (Lee et al. 2013).

It would be helpful when evaluating characteristics of patients with PHOMS to understand its pathogenesis and to confirm the relationship between PHOMS and ODD. This study aimed to evaluate the characteristics of and risk factors associated with PHOMS in children.

## Patients and Methods

This retrospective study was conducted according to the Declaration of Helsinki, after approval by the Institutional Review Board of the Nowon Eulji Medical Center.

The children under age 20 who were diagnosed with PHOMS between November 2015 and August 2018 at Eulji University Nowon Eulji Medical Center were reviewed (PHOMS group). The control group consisted of healthy children who underwent enhanced depth image (EDI) spectral-domain OCT without history of ophthalmologic surgery, neurologic and other ophthalmologic disease except refractive errors.

All children underwent a comprehensive ophthalmic examination that included a measurement of best-corrected visual acuity (BCVA), slit-lamp biomicroscopy, cycloplegic refraction (CR), ocular alignment test, dilated fundus examination, and color fundus photography. EDI spectral-domain OCT (Spectralis; Heidelberg Engineering, Dossenheim, Germany) was performed with 24-line radial scan images. All scans were reviewed and evaluated for the absence of motion artifacts and good centering on the optic discs. Optic nerve head diameters were defined as the Bruch’s membrane opening (BMO) and measured using the built-in measurement tool of the OCT machine. The mean values of the horizontal and vertical diameters of each plane were used. We also examined the thickness of macular ganglion cell layer (GCL) in the PHOMS group. The thickness values of GCL were measured in the Early Treatment Diabetic Retinopathy Study (ETDRS) central circular 3 mm diameter area including superior, inferior, temporal, and nasal areas (Yoo et al. 2019). If a patient in the PHOMS group was suspected to have concomitant true disc edema or mimicking papilledema, further follow-up examinations were performed to rule out other causes of disc swelling and to reveal any functional abnormalities. Additional testing may have included: Ishihara color vision test, fluorescein angiography, B-scan, static automated perimetry using a central 30-2 Humphry Field Analyzer (Humphrey Instruments Inc., San Leandro, CA), full-field visual evoked potentials, and brain magnetic resonance imaging. The spherical equivalent (SE) refractive error was calculated as the sphere + 1/2 cylinder determined by CR.

### Statistical analysis

Statistical analyses were performed using a commercially available statistical package (SPSS ver. 23.0 for Windows; IBM Inc., Armonk, NY). Continuous data are presented as a mean with standard deviation (SD), and categorical data are presented as counts and percentages. Univariable and multivariable logistic regression analyses were performed to investigate risk factors associated with the presence of PHOMS. In subgroup analysis, Fisher’s exact test was used to compare categorical data, and independent *t*-test was used for comparison of continuous parameters. A value of *P* < 0.05 was considered statistically significant.

## Results

### Baseline characteristics

A total of 112 children, 66 children with PHOMS (PHOMS group) and 46 children without PHOMS (control group) were analyzed in the study. Among the 66 children with PHOMS, 53 patients (80.3%) had bilateral POHMS and 13 patients (19.7%) had unilateral PHOMS. PHOMS occurred about equally in the right (60) and left (59) eyes. In the PHOMS group, there was no evidence of complications such as disc hemorrhage, nonarteritic anterior ischemic optic neuropathy, retinal vascular occlusion, or choroidal neovascular membrane. None of the patients had concomitant superficial ODD.

Patient demographics and characteristics are listed in Table 1. The mean age of the PHOMS group was 11.7 ± 2.6 years (range: 7 to 19) and 11.4 ± 3.1 years (range: 7 to 19) in the control group. Half of each group was male. All eyes in both groups had BCVA of 20/25 or greater. Mean SE was −3.13 ± 1.87 diopters (D) (range: −8.50 to +1.00) in the PHOMS group and −0.95 ± 2.65 D (range: −6.75 to +6.00) in the control group. Mean astigmatism was 0.67 ± 0.89 D (range: 0 to 4.50) and 0.88 ± 1.02 D (range: 0 to 3.50) in PHOMS and control groups, respectively. Mean disc size was 1735 ± 153 μm (range: 1347 to 2033) in the PHOMS group and 1741 ± 190 μm (range: 1296 to 2404) in the control group. All eyes in PHOMS group had myopia of −0.50 D or less, except an eye with SE +1.00 D. In Figure 1 and 2, we present cases of developing PHOMS with a concurrent myopic shift.

**Table 1.**
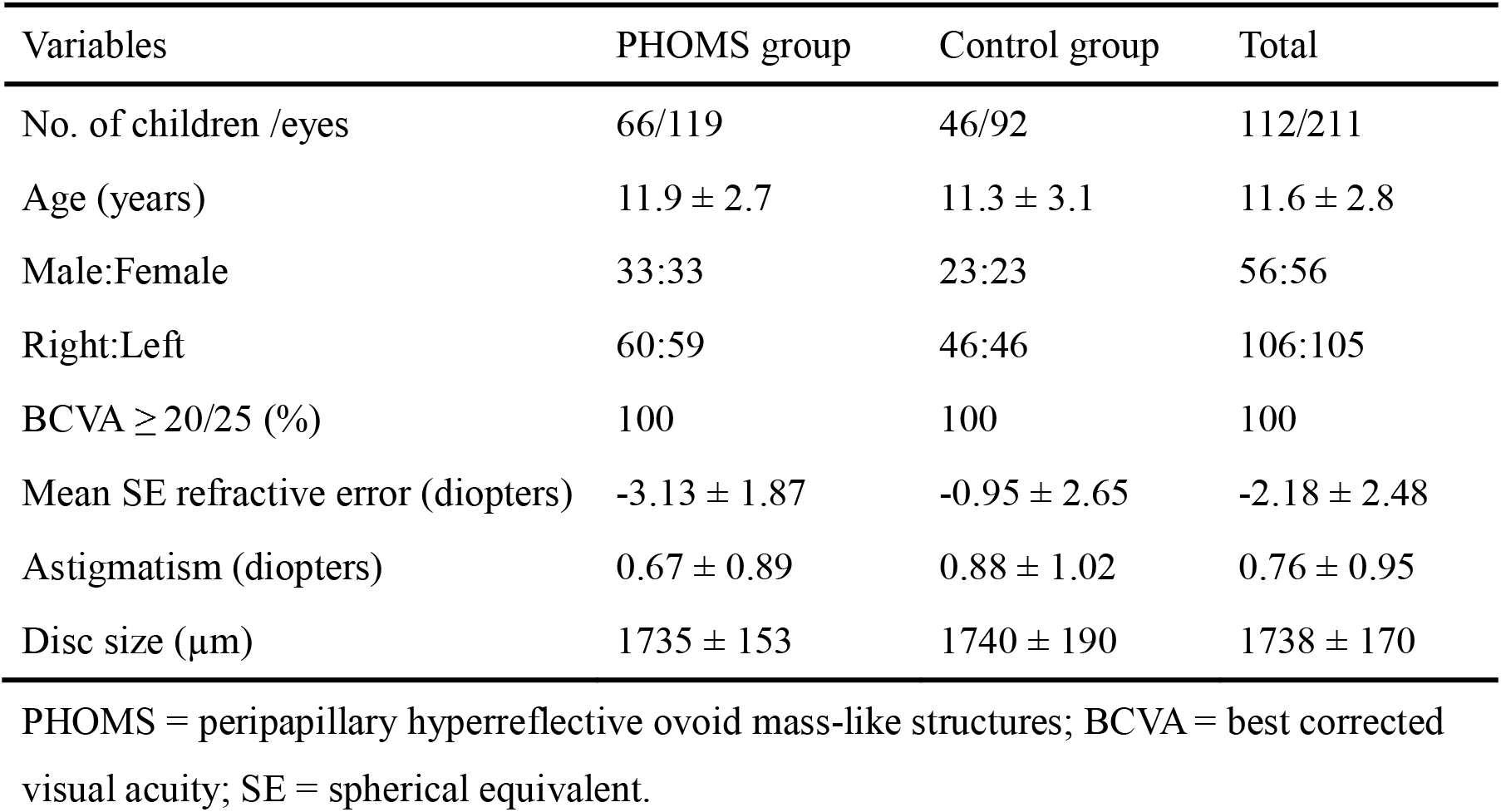
Demographics and characteristics of peripapillary hyperreflective ovoid mass-like structures (PHOMS) group and control group.

**Fig. 1.**
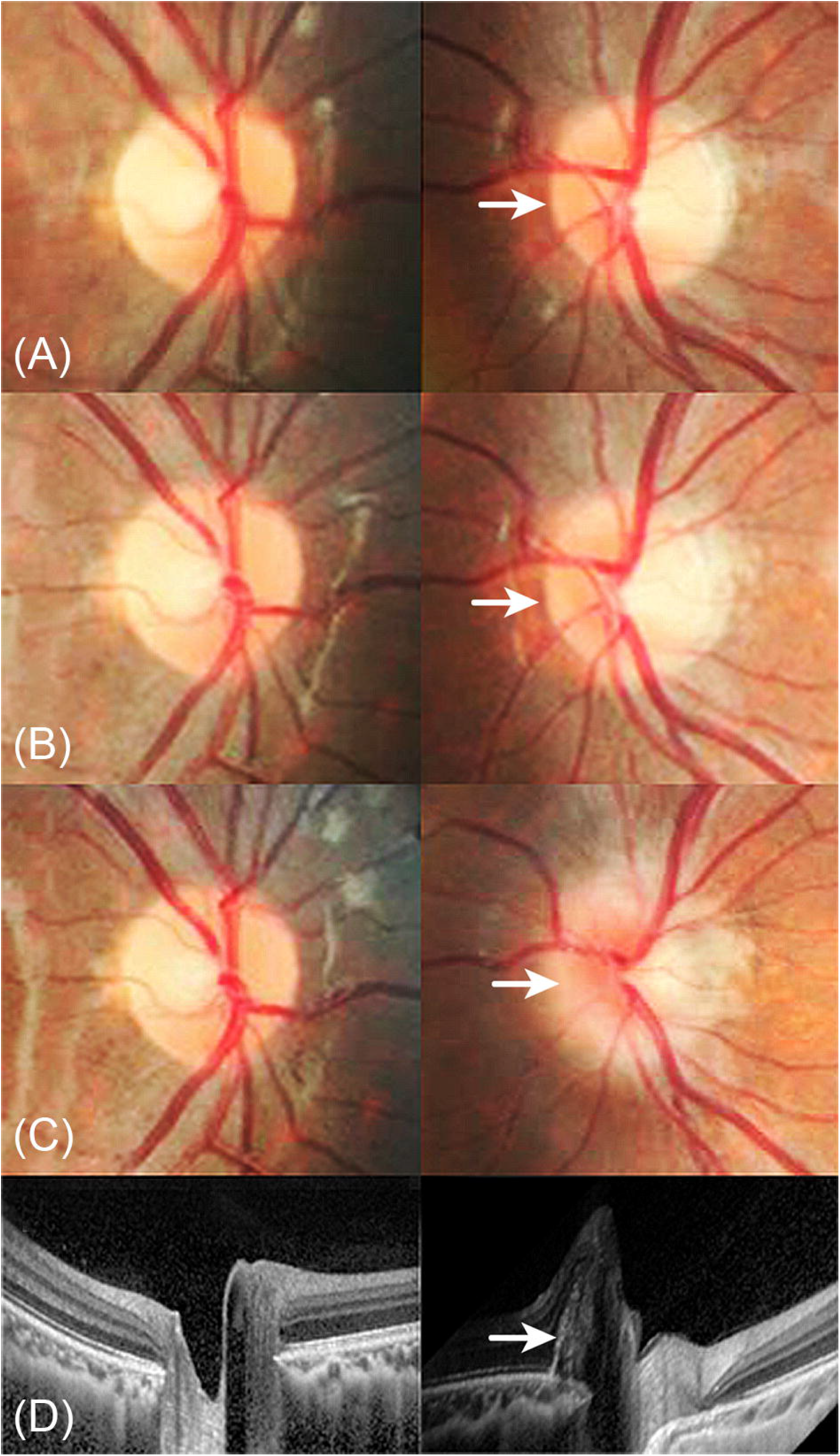
Serial changes (ages 8 to 11) of disc photographs and refractive errors in a male diagnosed with peripapillary hyperreflective ovoid mass-like structures (PHOMS). A myopic shift occurred in the left eye with the development of PHOMS. (**A**) The optic disc seems normal in both eyes at age 8. The spherical equivalent (SE) was +1.10 diopters (D) in the right eye and −0.50 D in the left eye. (**B**) The left disc margin was slightly elevated after 1.5 years. The SE was +1.10 D in the right eye and −1.80 D in the left eye. (**C**) Left disc blurring was aggravated at age 11 with SE +1.10 D in the right eye and −2.50 D in the left eye. (**D**) An EDI-optical coherence tomography (OCT) image showing PHOMS in the left eye at age 11.

**Fig. 2.**
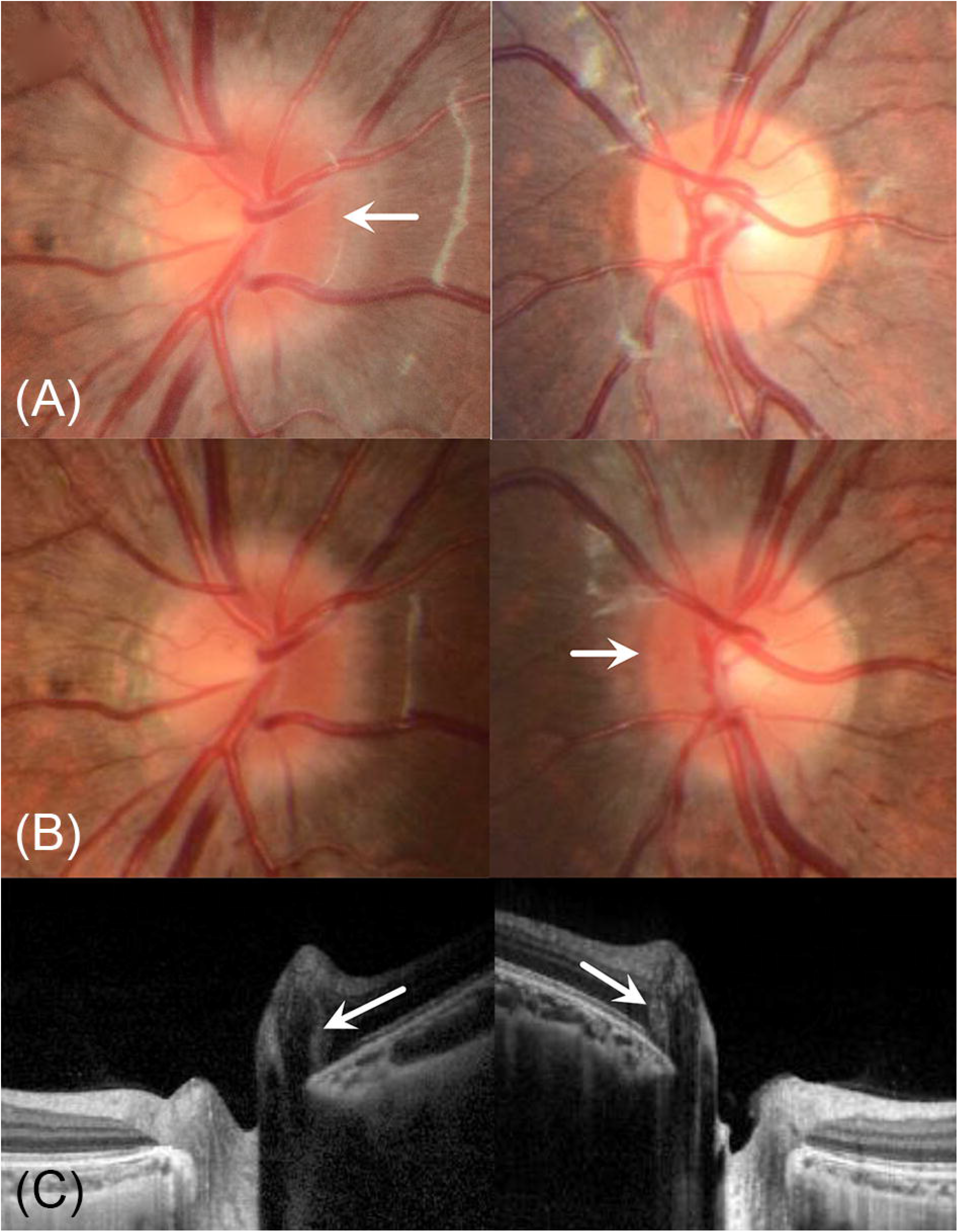
Another case of serial changes of peripapillary hyperreflective ovoid mass-like structures (PHOMS). The patient had follow-up for orthokeratologic lens in right eye. (**A**) Disc photographs at age of 9, the SE was −1.75 D in the right eye and −0.25 D in the left eye. The right nasal disc margin was blurred and left disc looks normal. (**B**) At age of 11, the SE was −1.75 D in the right eye and −0.75 D in the left eye. Along with left myopic shift, a new marginal blurring has been detected in left eye. (**C**) An EDI-optical coherence tomography (OCT) image showing small sized PHOMS as well as right eye, at this time.

### Risk factors associated with PHOMS

According to the univariable logistic analysis, SE decreased by 1D was significantly associated with PHOMS (odds ratio [OR] 1.59; confidential interval [CI] 1.35-1.86; *P* < 0.001) and this statistical significance remained after multiple logistic analyses (OR 2.0; CI 1.47-2.72; *P* < 0.001). We found no statistically significant difference between the two groups with respect to age, sex, laterality, astigmatism, and disc size (Table 2).

**Table 2.** Risk factors for peripapillary hyperreflective ovoid mass-like structures.

| Variables | Univariable model |  |  | Multivariable model* |  |  |
| --- | --- | --- | --- | --- | --- | --- |
|  | OR | 95% CI | p-value | OR | 95% CI | p-value |
| Age<br>per 1 year older | 0.95 | 0.83-1.09 | 0.493 |  |  |  |
| Male sex | 1.00 | 0.47-2.12 | 1.000 |  |  |  |
| Right eye | 0.98 | 0.57-1.70 | 0.952 |  |  |  |
| Mean SE refractive error<br>per 1 diopter decrease | 1.59 | 1.35-1.86 | <0.001 | 2.00 | 1.47-2.72 | <0.001 |
| Astigmatism<br>per 1 diopter increase | 1.26 | 0.94-1.68 | 0.124 |  |  |  |
| Disc size<br>per 100 μm increase | 1.00 | 0.99-1.01 | 0.812 | | | |

OR = odds ratio; CI = confidence interval; SE = spherical equivalent.
\*all variables were adjusted.

### Ganglion cell layer changes in PHOMS

We also analyzed the GCL thickness in patients with PHOMS via OCT. GCL thickness was slightly decreased in all sectors (temporal, nasal, superior, inferior, and average) in affected eyes compared with unaffected eyes; however, the difference was not statistically significant in any sector (Table 3).

**Table 3.** Comparison of macular ganglion cell layer thickness in affected eyes and unaffected eyes in patients with peripapillary hyperreflective ovoid mass-like structures.

| Variables | Affected eyes (N=119) | Unaffected eyes (N=13) | p-value |
| --- | --- | --- | --- |
| GCL temporal | 49.5 | 50.4 | 0.47 |
| GCL superior | 53.5 | 54.0 | 0.62 |
| GCL nasal | 52.2 | 54.3 | 0.77 |
| GCL inferior | 53.1 | 53.8 | 0.32 |
| GCL average | 52.1 | 53.1 | 0.28 |

GCL = ganglion cell layer.

### Subgroup analysis of unilateral PHOMS

In a subgroup analysis of 13 patients with unilateral PHOMS, the mean SE in the affected eye (−3.08 ± 1.77 D) was greater than in the fellow eye (−1.34 ± 2.26 D). This difference was statistically significant (*P* = 0.039) (Table 4). In the all cases of unilateral PHOMS, the affected eye was more myopic by at least −0.50 D than the fellow eye.

**Table 4.** Comparison of affected eyes and fellow eyes in cases with unilateral peripapillary hyperreflective ovoid mass-like structures (PHOMS) (N=13)

| Variables | Affected eyes | Fellow eyes | p-value |
| --- | --- | --- | --- |
| Mean SE refractive error (D) | -3.08 ± 1.77 | -1.34 ± 2.26 | **0.039** |
| Astigmatism (D) | 0.46 ± 0.71 | 0.71 ± 0.82 | 0.446 |
| Disc size (µm) | 1696 ± 128 | 1700 ± 212 | 0.961 |
| Right:Left | 7:6 | 6:7 | 1.000* |

SE = spherical equivalent; D = diopters.
\*Fischer's exact test

## Discussion

Peripapillary hyperreflective ovoid mass-like structures have been distinguished in recent years as an independent diagnosis (Malmqvist et al. 2018b) In previous studies, PHOMS was diagnosed as buried drusen or type 2 ODD (Lee et al. 2011; Gili et al. 2013; Lee et al. 2013; Bassi & Mohana 2014; Lee et al. 2018a). There is ongoing discussion about the diagnoses of PHOMS and buried drusen (Lee et al. 2018b; Malmqvist et al. 2018a, b) and its pathogenesis.

We compared the children with PHOMS to controls and found that myopia is a risk factor for PHOMS. Both univariable and multivariable models showed that when SE is decreased by 1 D, the risk of PHOMS is increased by about 2 times (*P* <0.001). All eyes in the PHOMS group had myopia of −0.50 D or less, except an eye with +1.00 D of hyperopia. The subgroup analysis of patients with unilateral PHOMS also supported this result. Myopia of the affected eyes was significantly greater than in fellow eyes (−3.08 ± 1.77 D and −1.34 ± 2.26 D, respectively). We identified cases of developing PHOMS with a myopic shift, documenting the changes with serial optic disc photographs.

Although herniation of distended axons into the peripapillary retina and exoplasmic stasis are suggested pathophysiologic causes of PHOMS (Malmqvist et al. 2018a, b), the exact pathophysiology of PHOMS is still unknown. We hypothesize that a myopic shift during adolescence is associated with the genesis of PHOMS and that optic disc tilt may be a mediator between myopia and PHOMS. Optic disc tilt is a feature of myopic shift, arising from scleral stretching in childhood (Kim et al. 2012). Nasal bulging and kinking of retinal nerve fibers might develop with the process of disc tilt during myopic shifts. Optic disc tilting also leads to the compression of axons and alterations of axonal transport on the nasal side of the optic nerve head, both of which are potential pathogenic causes of PHOMS (Tso 1981).

Our theory is supported by the findings in several studies of tilted disc syndrome (Shinohara et al. 2013; Pichi et al. 2014). Shinohara et al. and Pichi et al. evaluated morphology of tilted discs using OCT. They reported the peripapillary herniation of retinal nerve fiber inside Bruch’s membrane in patients with tilted disc syndrome, though they did not use the terminology “PHOMS” (Shinohara et al. 2013; Pichi et al. 2014). Seo and Park described a case of rapidly progressing PHOMS in a 9-year-old child, consistent with the age range in which a myopic shift usually acquired (Seo du & Park 2015).

We also found several studies presenting the emergence and progression of superficial ODD in childhood. Giuffre reported 2 cases of ODD and suggested that disc tilt and ODD can have a cause-effect relationship due to axonal compression induced by distortion of the scleral canal in tilted discs (Giuffre 2005). Frisén reported a case of ODD that was followed over 23 years. In this case report, drusen showed dynamic morphologic changes from the ages of 8 to 16, again an age range in which myopia usually progresses (Frisen 2008). Malmqvist et al. followed 8 patients with superficial ODD over 56 years and concluded that the progression of ODD might occur before adulthood (Malmqvist et al. 2017). This commonality between PHOMS and ODD is probably caused by the shared pathogenesis of compression of axons and axonal stasis.

There are ongoing debates about the diagnosis of PHOMS vs. buried drusen, and whether PHOMS are early ODD (Lee et al. 2018b; Malmqvist et al. 2018a). Whether subretinal hyperreflective masses on OCT are diagnosed as PHOMS or buried drusen, our hypothesis remains valid because optic disc tilting leads to the compression of axons and alterations of axonal transport on the nasal side of the optic nerve head, which is considered the most likely pathogenesis of both conditions (Tso 1981). Further prospective longitudinal studies are needed to better distinguish PHOMS and ODD.

Small disc size (Lee et al. 2013; Ghassibi et al. 2017; Malmqvist et al. 2018c) has been reported as a risk factor for buried ODD/PHOMS. However, in our study, the disc sizes were similar in the PHOMS and control groups, and the disc size as measured by BMO was not related to the presence of PHOMS (*P =* 0.812). Lee et al. found that patients with buried drusen had smaller disc sizes than patients with superficial drusen, and that smaller disc size is associated with larger sized buried drusen (Lee et al. 2013). The buried drusen in this study were consistent with PHOMS. However, the characteristics of these two groups of patients were significantly different. The mean age of the visible drusen group was about 53 and for the PHOMS group, it was 13 (Lee et al. 2013). A report from the Copenhagen Child Cohort Eye Study indicated that the disc size in ODD patients was significantly smaller than that in controls (Malmqvist et al. 2018c). ODD was defined in this study as “circumscribed hyporeflective elements, fully or partially surrounded by a hyporeflective border”, not the same definition as PHOMS. The discrepancy in the results regarding disc size may be attributable to the use of different patient populations and different definitions of ODD/PHOMS between the studies. Nevertheless, we impressed that larger optic nerve heads are less likely to develop PHOMS in spite of myopic progression. None of the patients having a disc diameter greater than 2033 μm showed PHOMS in our study. Again, further prospective longitudinal studies are needed to better characterize the relationship between optic disc size and risk of PHOMS.

In the study of PHOMS mentioned above, Lee et al. speculated that factors other than small disc size might be related to PHOMS genesis, because they found that disc sizes in affected eyes were larger than in the unaffected corresponding eye in patients with unilateral PHOMS (Lee et al. 2013). Our finding that myopia is a risk factor for PHOMS may provide further insight into understanding the genesis of PHOMS.

There are several limitations to this study. First, this study is a retrospective cross sectional study. Second, axial length was not evaluated, even though myopia is usually correlated with the elongation of ocular axial length. Therefore, further larger prospective longitudinal studies are needed to support our hypothesis and to gain a better understanding of PHOMS. Despite these limitations, this is the first reported association between myopia and PHOMS.

According to our findings, myopia increases the risk of PHOMS in children. Disc tilt induced by myopic shifts during childhood may be associated with PHOMS. These findings might increase the understanding of the genesis of PHOMS.

## Data Availability

Data are available upon request.

## Author’s contributions

study design (I.J.L. and S.Y.O.); conduction of the study (I.J.L.); data collection and management (I.J.L.); data analysis and interpretation (I.J.L., K.A.P.); drafting the manuscript (I.J.L.) and the review and final approval of the manuscript (I.J.L., K.A.P., S.Y.O.). All authors contributed and approved the final manuscript.

## References

Auw-Haedrich C, Staubach F & Witschel H (2002): Optic disk drusen. Surv Ophthalmol 47: 515–532.

Bassi ST & Mohana KP (2014): Optical coherence tomography in papilledema and pseudopapilledema with and without optic nerve head drusen. Indian J Ophthalmol 62: 1146–1151.

Casado A, Rebolleda G, Guerrero L, Leal M, Contreras I, Oblanca N & Munoz-Negrete FJ (2014): Measurement of retinal nerve fiber layer and macular ganglion cell-inner plexiform layer with spectral-domain optical coherence tomography in patients with optic nerve head drusen. Graefes Arch Clin Exp Ophthalmol 252: 1653–1660.

Friedman AH, Beckerman B, Gold DH, Walsh JB & Gartner S (1977): Drusen of the optic disc. Surv Ophthalmol 21: 373–390.

Frisen L (2008): Evolution of drusen of the optic nerve head over 23 years. Acta Ophthalmol 86: 111–112.

Ghassibi MP, Chien JL, Abumasmah RK, Liebmann JM, Ritch R & Park SC (2017): Optic Nerve Head Drusen Prevalence and Associated Factors in Clinically Normal Subjects Measured Using Optical Coherence Tomography. Ophthalmology 124: 320–325.

Gili P, Flores-Rodriguez P, Yanguela J, Orduna-Azcona J & Martin-Rios MD (2013): Sensitivity and specificity of monochromatic photography of the ocular fundus in differentiating optic nerve head drusen and optic disc oedema: optic disc drusen and oedema. Graefes Arch Clin Exp Ophthalmol 251: 923–928.

Giuffre G (2005): Optic disc drusen in tilted disc. Eur J Ophthalmol 15: 647–651.

Kim TW, Kim M, Weinreb RN, Woo SJ, Park KH & Hwang JM (2012): Optic disc change with incipient myopia of childhood. Ophthalmology 119: 21-26 e21-23.

Lee KM, Woo SJ & Hwang JM (2011): Differentiation of optic nerve head drusen and optic disc edema with spectral-domain optical coherence tomography. Ophthalmology 118: 971–977.

Lee KM, Woo SJ & Hwang JM (2013): Morphologic characteristics of optic nerve head drusen on spectral-domain optical coherence tomography. Am J Ophthalmol 155: 1139–1147 e1131.

Lee KM, Woo SJ & Hwang JM (2018a): Factors associated with visual field defects of optic disc drusen. PLoS One 13: e0196001.

Lee KM, Woo SJ & Hwang JM (2018b): Peripapillary Hyperreflective Ovoid Mass-Like Structures: Is It Optic Disc Drusen or Not? J Neuroophthalmol 38: 567–568.

Malmqvist L, Bursztyn L, Costello F et al. (2018a): Peripapillary Hyperreflective Ovoid Mass-Like Structures: Is It Optic Disc Drusen or Not?: Response. J Neuroophthalmol 38: 568–570.

Malmqvist L, Bursztyn L, Costello F et al. (2018b): The Optic Disc Drusen Studies Consortium Recommendations for Diagnosis of Optic Disc Drusen Using Optical Coherence Tomography. J Neuroophthalmol 38: 299–307.

Malmqvist L, Li XQ, Eckmann CL, Skovgaard AM, Olsen EM, Larsen M, Munch IC & Hamann S (2018c): Optic Disc Drusen in Children: The Copenhagen Child Cohort 2000 Eye Study. J Neuroophthalmol 38: 140–146.

Malmqvist L, Lund-Andersen H & Hamann S (2017): Long-term evolution of superficial optic disc drusen. Acta Ophthalmol 95: 352–356.

Pichi F, Romano S, Villani E et al. (2014): Spectral-domain optical coherence tomography findings in pediatric tilted disc syndrome. Graefes Arch Clin Exp Ophthalmol 252: 1661–1667.

Pollack IP & Becker B (1962): Hyaline bodies (drusen) of the optic nerve. Am J Ophthalmol 54: 651–654.

Seo du R & Park SH (2015): Case of rapidly progressing buried optic nerve head drusen. JAMA Ophthalmol 133: e143467.

Shinohara K, Moriyama M, Shimada N, Nagaoka N, Ishibashi T, Tokoro T & Ohno-Matsui K (2013): Analyses of shape of eyes and structure of optic nerves in eyes with tilted disc syndrome by swept-source optical coherence tomography and three-dimensional magnetic resonance imaging. Eye (Lond) 27: 1233-1241; quiz 1242.

Tso MO (1981): Pathology and pathogenesis of drusen of the optic nervehead. Ophthalmology 88: 1066–1080.

Yoo YJ, Hwang JM & Yang HK (2019): Inner macular layer thickness by spectral domain optical coherence tomography in children and adults: a hospital-based study. Br J Ophthalmol.

